# How lifestyle changes within the COVID-19 global pandemic have affected the pattern and symptoms of the menstrual cycle

**DOI:** 10.1101/2021.02.01.21250919

**Authors:** Georgie Bruinvels, Esther Goldsmith, Richard C. Blagrove, Dan Martin, Laurence Shaw, Jessica Piasecki

**Affiliations:** Orreco Ltd, National University of Ireland Business, Innovation Centre, Galway, H91 RW53, Ireland; School of Sport, Exercise and Health Sciences, Loughborough University, Loughborough, LE11 3TU, UK; School of Sport and Exercise Sciences, University of Lincoln, Lincoln, LN6 7GA, UK; School of Science and Technology, Nottingham Trent University, Nottingham, NG11 8NS, UK

**Keywords:** COVID-19, exercise, females, menstrual cycle, nutrition

## Abstract

**Background:** The coronavirus 2019 (COVID-19) pandemic has caused significant changes to homes, working life and stress. The purpose of this research was to investigate the implications that the COVID-19 pandemic has had on the menstrual cycle and any contributing factors to these changes.

**Methods:** A questionnaire was completed by 749 participants, whom ranged from ‘physically active’ to elite, in their training status. The questionnaire captured detail on menstrual cycle symptoms and characteristics prior to and during the COVID-19 pandemic lockdown period, as well as lifestyle, stress, exercise and nutrition. Descriptive statistics and frequency distribution were reported and decision tree analysis performed. Statistical significance was assumed at p<0.05.

**Results:** Fifty-two point six percent of females experienced a change in their menstrual cycle during the lockdown period. Psychosocial symptoms had changed in over half of all participants. Participants who reported increased stress/worry in family and personal health were significantly associated with changes in menstrual symptoms. Similarly, job security stress was associated with increases in bleeding time (p<0.05).

**Conclusions:** It is important that females and practitioners become aware of the implications of stressful environments and the possible long-term implications on fertility, particularly given the uncertainty around a second wave of the global pandemic.

## 1. Introduction

The menstrual cycle is a natural process for most females that occurs in the reproductive years between puberty and menopause. A ‘normal’ (eumenorrheic) menstrual cycle lasts 22-35 days (Fehring, Schneider and Raviele, 2006) and is characterised by a cyclical fluctuation of sex hormones mediated by the hypothalamic-pituitary axis, which is essential for maintenance of bone health and fertility. Whilst oestrogen and progesterone are the key sex hormones involved in the reproductive system, they are also vital in the regulation of other physiological systems and maintaining holistic health. In particular, oestrogen is a key regulator of bone resorption (Chen, Wang and Huang, 2009) and also has a cardioprotective role (Iorga *et al*., 2017). Therefore, continuous oestrogen exposure through the menstrual cycle post-puberty and pre-menopause could also reduce risk of other health conditions such as osteoporosis and cardiovascular disease (Bae, Park and Kwon, 2018; Mumford *et al*., 2012).

A eumenorrheic menstrual cycle can be used to indicate that the body is in an adaptive physiological state, and not subject to excess stress. Therefore, from a physical performance perspective, it can be used to guide readiness and adaptation. Dysregulation of this process may be multifactorial; often occurring as a result of perturbation of the hypothalamic-pituitary axis (Roca *et al*., 2003), which causes a reduction in the secretion and production of ovarian steroids (Nappi *et al*., 1993). Irregularities often present with anovulation, and may involve changes in cycle length or the cessation of cycles (hypothalamic amenorrhea) (Bae, Park and Kwon, 2018; Kato *et al*., 1999). Whilst prolonged energy deficiencies are often cited as being the aetiology of functional hypothalamic amenorrhea (De Souza *et al*., 2020), other causes have been reported, such as sleep disturbances, lifestyle stress and alterations in circadian timing systems (Lateef and Akintubosun, 2020). Up to 90% of women of a reproductive age also experience menstrual cycle symptoms (Halbreich, 2003), and severe symptoms have consistently been shown to affect quality of life (Choi *et al*., 2010; Kahyaoglu Sut and Mestogullari, 2016) as well as negatively impacting exercise performance (Findlay *et al*., 2020; Chantler, Mitchell and Fuller, 2009). Premenstrual symptoms are positively associated with poor diet and lack of exercise (Rad, Sabzevary and Dehnavi, 2018; Cheng *et al*., 2013), as well as increased alcohol intake (Fernández *et al*., 2018).

Psychological stress has frequently been associated with alterations to menstrual cycle duration and the number and severity of symptoms (Nagma *et al*., 2015; Palm-Fischbacher and Ehlert, 2014); illustrated by the increase in the prevalence of amenorrhea during wartime (DREW, 1961; Barsom *et al*., 2004). Recent research has reported that students under a high amount of perceived stress were four times more likely to be amenorrhoeic (Rafique and Al-Sheikh, 2018). Furthermore, women with stressful jobs were twice as likely to experience a decreased cycle length (Fenster *et al*., 1999), often as a consequence of decreased follicular phase length, with little variation in the length of the luteal phase (McIntosh *et al*., 1980). However, stressful environments have not consistently been shown to cause changes in the length or regularity of the menstrual cycle characteristics (Williams *et al*., 2020; Clarvit, 1988; Nagata *et al*., 1986). Contradictions between findings may be due to individual variation in dispositional resilience, which may be a protective psychological trait regarding menstrual function in high stress scenarios (Palm-Fischbacher and Ehlert, 2014).

Coronavirus disease 2019 (COVID-19) is a highly contagious and, for some, a fatal disease. The spread of COVID-19 has resulted in a global pandemic and public health emergency. The pandemic has had a widespread, negative impact on many aspects of society, and there is growing concern that necessary measures such as self-isolation and limits on exercise and travel may negatively impact psychological health (Mehrsafar *et al*., 2020; World Health Organisation, 2020). The severe acute respiratory syndrome and middle-east respiratory syndrome epidemics have both previously demonstrated increases in psychiatric comorbidities such as depression, anxiety, panic attacks and suicidality (Cheung, Chau and Yip, 2008; Mak *et al*., 2009; Zhang *et al*., 2020b). Recent surveys in China following the onset of COVID-19 have reported that 25% of college students experienced symptoms of anxiety, whilst 53% of Chinese residents deemed the psychological impact of COVID-19 to be moderate or severe (Wang *et al*., 2020; Cao *et al*., 2020). Recent research has identified reduced sleep quality during the COVID-19 outbreak as the primary mediator of negative emotions (e.g. stress and anxiety) during this time (Zhang *et al*., 2020b). However, the authors did find that regular exercise could offset this (Zhang *et al*., 2020b). There is little evidence concerning the effect of living through a pandemic or epidemic on menstrual function. This is partly due to the limited representation of female participants in research following these crises (Wenham *et al*., 2020). However, following the 2014-2016 West African Ebola virus epidemic, one study conducted in Liberia found that 27% women who had been eumenorrheic before the outbreak, experienced irregular menstruation afterwards (Godwin *et al*., 2019). However, as the Ebola virus also resulted in rapid weight loss, the authors note that it is unclear the specific reasons for which the irregularities occurred.

COVID-19 has resulted in significant upheaval of daily life for individuals worldwide, with changes to work, social, diet, exercise patterns (Ammar *et al*., 2020; Ashby, 2020; Zhang *et al*., 2020a) which has resulted in increased stress (Ozamiz-Etxebarria *et al*., 2020; Torales *et al*., 2020; Vindegaard and Benros, 2020). For exercising females there is a greater risk of menstrual dysfunction (Baker, 1981; Slater *et al*., 2016), this combined with the heightened stress of COVID-19, could result in nationwide spread of exacerbated menstrual cycles symptoms and/or increased susceptibility to changes in cycle length. Given the importance of the menstrual cycle for females, the aim of this study was to investigate the different symptoms and alterations to the menstrual cycle that females may have experienced during the COVID-19 lockdown period, and the main factors that contribute to these disturbances.

## 2. Materials and Methods

### 2.1. Ethics and Participant recruitment

The study was conducted in accordance with the declaration of Helsinki and approved by the institutional ethics committee at Nottingham Trent University. Participant recruitment and data collection took place from 27^th^ May 2020 – 17^th^ June 2020. Participants were eligible to if they were; aged ≥18 years; exercising at any level; eumenorrheic (pre COVID-19) with or without a form of hormonal contraception. Participants must have experienced a minimum of nine menstrual cycles or withdrawal bleeds over the past 12 months (prior to COVID-19).

### 2.2. Survey Development

The questionnaire contained 33 questions and was developed by all researchers, whom collectively have extensive experience with questionnaire design. The survey platform Qualtrics (Qualtrics 2005; 37, 892 ed. Provo, Utah, USA) was used to make the questionnaire accessible online. The questionnaire was compiled of mainly binary or multiple-choice questions. The online link provided to the questionnaire began with the participant information sheet, followed by participant consent. By proceeding to the next stage participants were able to consent to the study. If participants did not wish to proceed, they were able to close the browser prior to consent. All participants provided themselves with a unique participant identification to ensure anonymity.

The survey comprised of five sections. The first section provided an introduction and contained demographic related questions such as age, occupation, primary sport, level of competition, menstrual status, cycle length prior to COVID-19, marital status and ethnicity. The second section captured detail about the participants’ working status prior to COVID-19 and during the COVID-19 pandemic lockdown, information around children and if applicable, how this affected their working routine. This section also included Likert scales to detail how financial status and job security affected worry and stress. The third section requested detail on exercise behaviour changes that may or may not have occurred for the participant during lockdown. There were ten different categories of training in which the participants could answer with one of the following options: 1; about the same, 2; increased, 3; decreased, 4; I never do this. The fourth section contained questions concerning the menstrual experiences of the participants, based on previous work by co-author’s xxx. The same options used in section three applied here for 28 different symptoms associated with the menstrual cycle. Participants were also asked about the length of their cycle and bleeding patterns in a multiple-choice format (increased, decreased, stayed the same), with an opportunity to elaborate on coping mechanisms for any symptoms that may have arisen during the COVID-19 lockdown. The final section of the questionnaire focused on the general health of the participant. This included Likert scales regarding worry around personal and family health, and whether the participants have been previously diagnosed with, or have been assumed to have/had, COVID-19. The final element of the survey captured aspects of nutritional intake that may have changed during the lockdown using the previously mentioned scale (1-4) from sections 3 and 4. Data processing is shown in Fig. 1.

**Figure 1.**
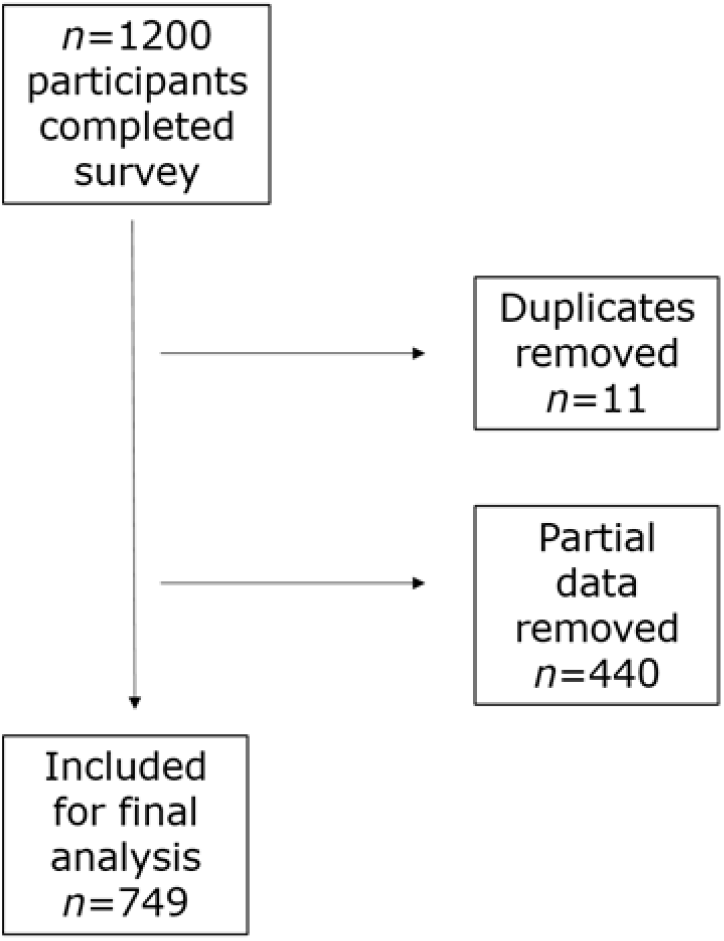
Flow diagram of data inclusion.

### 2.3. Data Analysis

Data were analysed using IBM SPSS (v. 23.0) and Microsoft Excel. Partial responses were removed from the dataset. Data was then coded to categorised responses and then frequency analysis was carried out. Data are represented as mean ± standard deviation (SD), frequencies and percentages. Statistical significance was set at p ≤ 0.05. Further analysis was carried out using R Statistical Software. Decision trees were created using the *rpart (Therneau and Atkinson, 2019)* and *rpart.plot (Milborrow, 2019)* packages to explore possible connections between changes in menstrual symptoms since the onset of COVID-19 and specific lifestyle changes covered in the questionnaire. These trees were pruned using 10-fold cross-validation to prevent over-fitting. Potential relationships between changes in symptoms and lifestyle identified by the decision trees were then tested using Fisher’s exact test.

## 3. Results

### 3.1 Participant Characteristics

Participant characteristics and demographics are displayed in Table 1. 97.2% of the sample did not have a diagnosis of COVID-19 at the time of reporting, with 13.3% declaring they had experienced some symptoms of COVID-19 and had therefore had to self-isolate. Ten percent reported that someone in their household had experienced symptoms and in turn they were required to self-isolate.

**Table 1.** Participant characteristics and demographics (n=749).

| Characteristic | Categories | % |
| --- | --- | --- |
| Age (years) | 15-20 | 5.1 |
|  | 21-30 | 41.0 |
|  | 31-40 | 34.9 |
|  | 41-50 | 18.0 |
|  | 51+ | 1.0 |
| Activity level | Active/fitness training (5 hrs or less per week) | 51.3 |
|  | Sub elite training (training towards a goal, 5-8 hrs per week) | 32.1 |
|  | Elite training (national/international competition, 8 hrs + per week) | 14.5 |
|  | Professional Athlete | 2.1 |
| Marital Status | Divorced | 1.3 |
|  | Living with partner | 26.3 |
|  | Single | 35.7 |
|  | Married | 33.3 |
|  | Widowed | 0.3 |
|  | Other | 3.1 |
| Ethnicity | Asian/Asian Black | 1.3 |
|  | Black/African/Caribbean/Black British | 0.7 |
|  | Mixed/Multiple ethnic groups | 3.4 |
|  | Rather not say | 0.3 |
|  | White | 92.6 |
|  | Other | 1.7 |
| Do you have children? | Yes | 23.2 |
|  | No | 76.8 |

### 3.2 Participant Stress and Employment

Prior to COVID-19 >75% of our sample were in full-time employment. During COVID-19 lockdown half of the respondents reported to be working full-time at home and 17% were working part-time at home. Only 54 participants indicated they had been furloughed at either 80% or 100% pay. Distribution of working status prior to and during COVID-19 is shown in Fig. 2.

**Figure 2.**
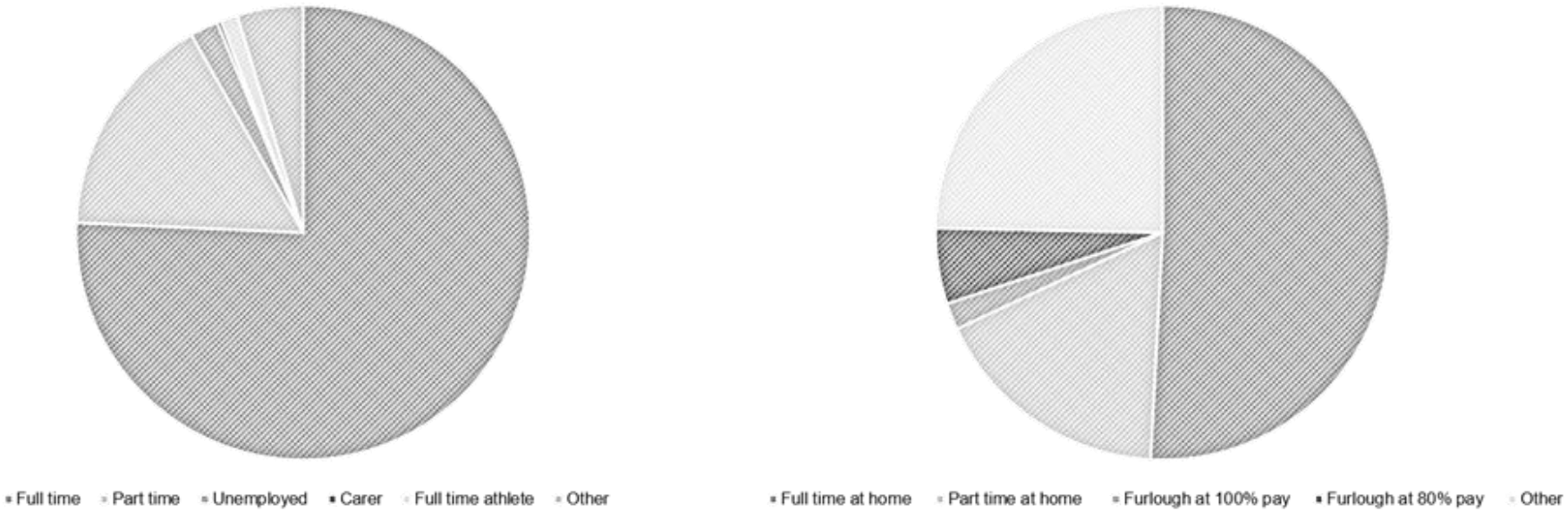
Working status prior to and during the COVID-19 pandemic. (Left) Working status prior to COVID-19; Full time 75.7%, part-time, 16.1%, unemployed 2.0%, Carer 0.3%, full time athlete 1.2%, other 4.7% (Right) Working status during COVID-19; full time at home 50.9%, part-time at home 17.2%, furlough at 100% 1.9%, furlough at 80& 5.4%, other 24.7%.

The stress experienced by participants is displayed in table 2. The majority of those with children expressed that their children caused higher levels of daily stress. When considering other possible sources of stress and worry such as financial stress, job security and stress regarding menstrual symptoms, most participants (>50%) did not report being overly worried or stressed (1-3 on Likert scale) by such parameters (Table 2).

**Table 2.** Likert stress scales.

| **Question** | **Likert Scale** | **Frequency** | **%** |
| --- | --- | --- | --- |
| How have children<br>affected daily stress | 1 | 11 | 6.5 |
|  | 2 | 17 | 10.0 |
|  | 3 | 24 | 14.1 |
|  | 4 | 40 | 23.5 |
|  | 5 | 50 | 29.4 |
|  | 6 | 19 | 11.2 |
|  | 7 | 8 | 4.7 |
| Worried about job<br>security | 1 | 254 | 34.1 |
|  | 2 | 125 | 16.8 |
|  | 3 | 90 | 12.1 |
|  | 4 | 70 | 9.4 |
|  | 5 | 63 | 8.5 |
|  | 6 | 49 | 6.6 |
|  | 7 | 42 | 5.6 |
| Worried about<br>personal finances | 1 | 210 | 28.2 |
|  | 2 | 131 | 17.6 |
|  | 3 | 117 | 15.7 |
|  | 4 | 86 | 11.5 |
|  | 5 | 68 | 9.1 |
|  | 6 | 42 | 5.6 |
|  | 7 | 44 | 5.9 |
| Stressed about<br>menstrual cycle<br>symptom changes | 1 | 193 | 25.9 |
|  | 2 | 114 | 15.3 |
|  | 3 | 84 | 11.3 |
|  | 4 | 68 | 9.1 |
|  | 5 | 49 | 6.6 |
|  | 6 | 26 | 3.5 |
|  | 7 | 21 | 2.8 |
| Worried about own<br>health | 1 | 166 | 22.3 |
|  | 2 | 171 | 23.0 |
|  | 3 | 137 | 18.4 |
|  | 4 | 113 | 15.2 |
|  | 5 | 84 | 11.3 |
|  | 6 | 32 | 4.3 |
|  | 7 | 23 | 3.1 |
| Worried about<br>Family health | 1 | 39 | 5.2 |
|  | 2 | 75 | 10.1 |
|  | 3 | 115 | 15.5 |
|  | 4 | 117 | 15.7 |
|  | 5 | 157 | 21.1 |
|  | 6 | 119 | 16.0 |
|  | 7 | 114 | 15.3 |

### 3.3 Menstrual Cycle Symptoms

Most changes to menstrual symptoms experienced during the COVID-19 pandemic were psychosocial in nature (Fig. 3). Over half of all participants reported to have experienced lack of motivation (59.3%), focus (53.1%) and concentration (54.9%). Similarly, symptoms of mood changes, irritability, emotional feeling, worry and being distracted were all reported in over half of all participants. There were minimal differences in this pattern when dividing participants between those who were hormonal contraceptive users or not.

**Figure 3.**
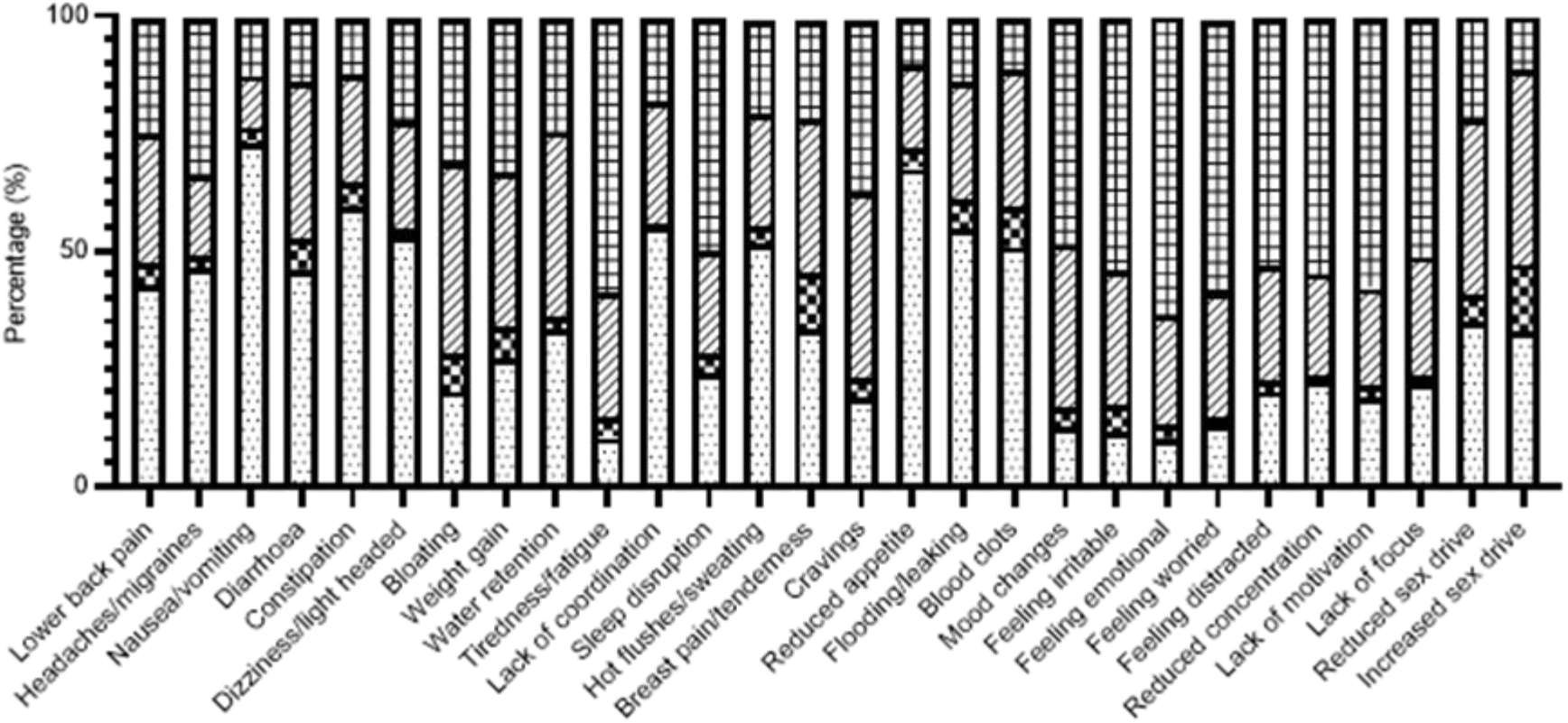
Percentage of symptoms reported by all participants. Squares indicate increase in symptoms, dashes represent stayed the same, dark squares represent decrease and dots represent do not experience this symptom.

Decision tree analysis indicated differences in the symptoms suffered by women training at sub-elite level and those who considered themselves physically active. Of those women who experienced increased bloating, 54% were sub-elite level competitors or above, compared to only 46% of those participating below this level (p=0.035). However, the tree (Supplementary file, Figure 4) also revealed that higher level competitors may have been less likely than other women to experience reduced sex drive, constipation, reduced focus, reduced motivation, feeling emotional and lower back pain.

### 3.4 Nutrition and Exercise Patterns

Changes in nutritional and exercise patterns can be found in Table 4. When assessing both nutritional changes and exercise behaviours according to any changes in menstrual cycle length, a consistent pattern remains whether the females expressed a change in menstrual cycle length or not.

**Table 4.**
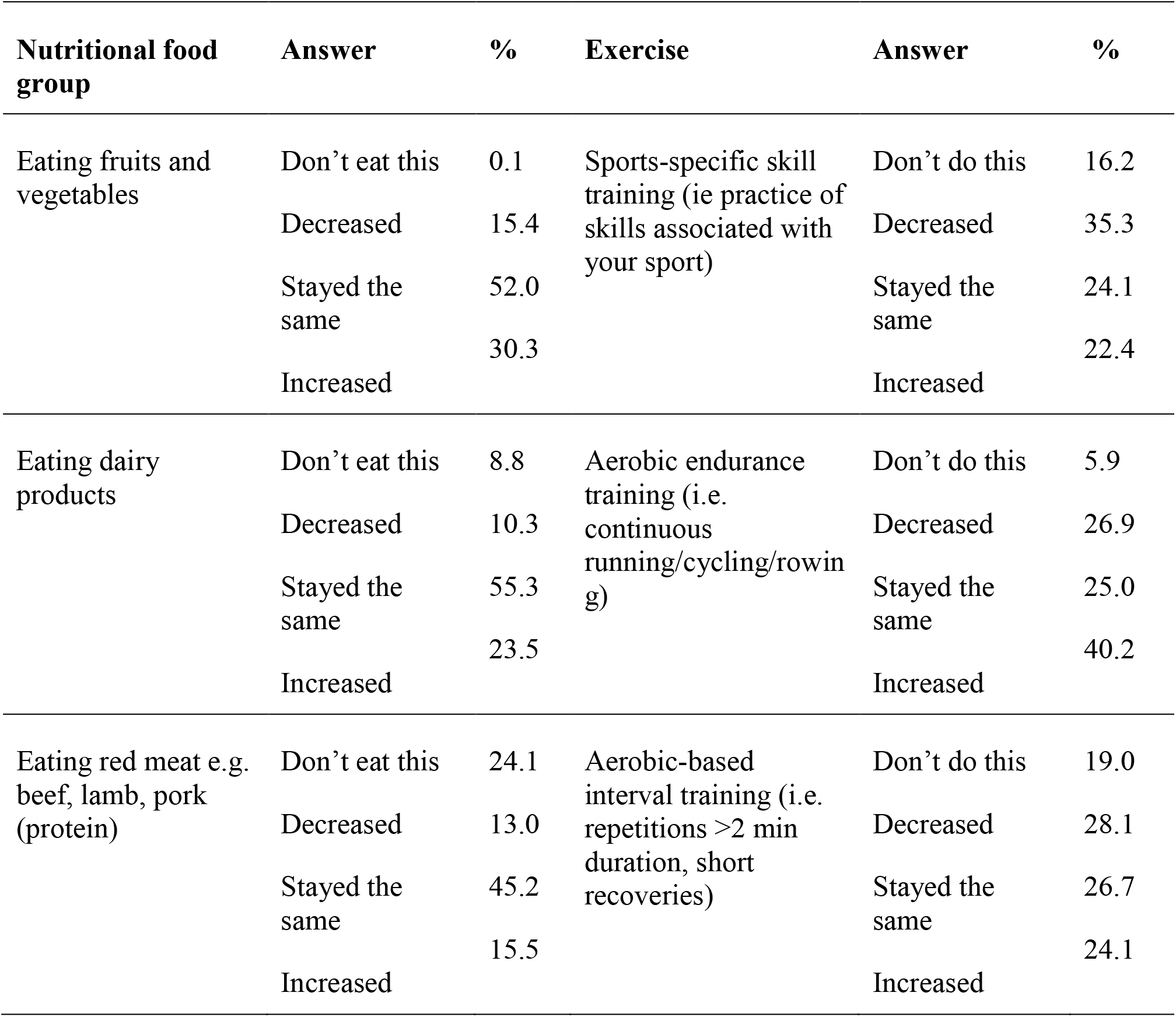
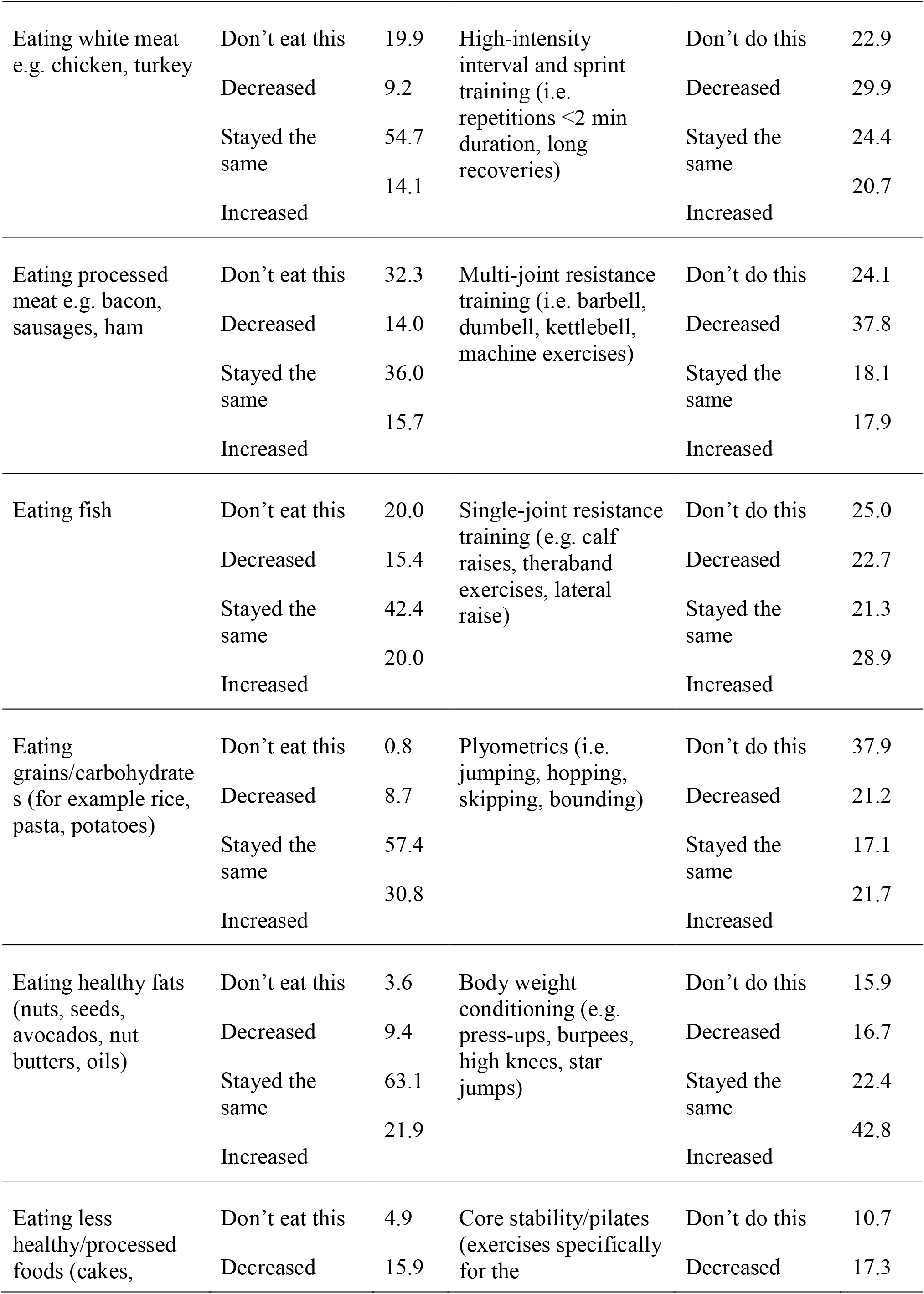
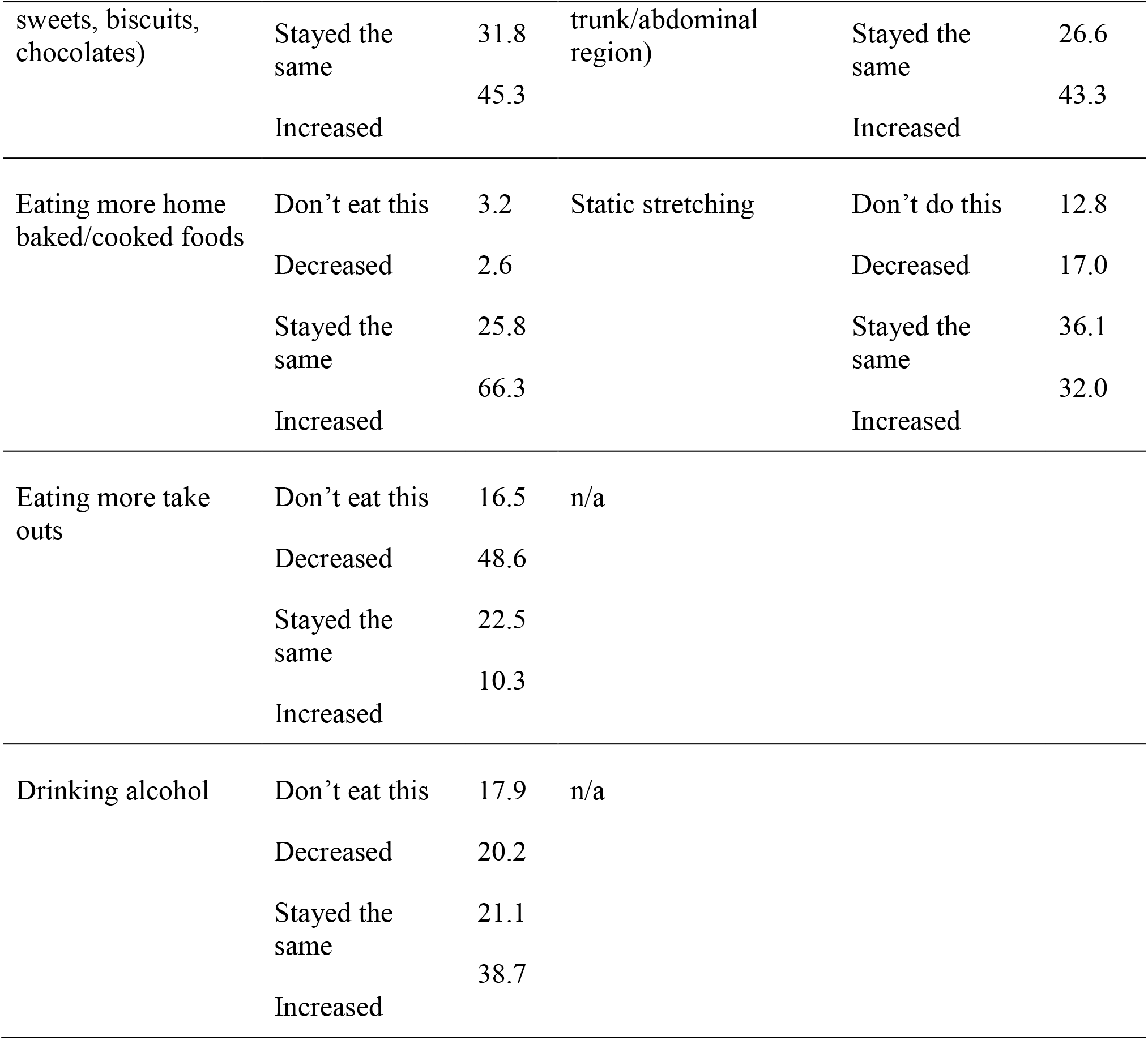
Nutritional and exercise behaviour changes for all female participants.

### 3.5 Decision Tree Analysis

Of the 28 symptoms listed in Fig. 3, women indicated a 9.4 point average increase during the pandemic (of the 28 symptoms given, the number represented here indicates the average number of symptoms that were reported to increase). However, decision tree analysis (Supplementary file, Figure 4) revealed that factors associated with ‘stress’ had the most significant impact on this increase in symptoms. Women who rated personal health stress pre-COVID-19 at three or higher (see Table 2) increased to 11 during the lockdown period and this score increased to 13 if additional stress concerning family health was given the highest rating. Among the women with lower levels of stress about personal health, the average increase in symptoms was 7.8 but this score only stayed low among women who maintained or increased their fruit and vegetable intake (7.3). Among those who did not, an average of 11 symptoms increased.

The cross-validated decision tree process did not identify any factors significantly related to blood-loss changes but did show that high levels of stress regarding job security (rating of 6 or 7) made it more likely that the time bleeding would change (Supplementary file, Figure 4). Of the 653 women not reporting high levels of job security stress, 34% experienced changes in time bleeding compared to 54% of the 91 women reporting high levels of stress. Fisher’s exact test suggested a significant relationship (p<.001). Of the 91 women reporting high levels of job security stress, 38 reported that they had maintained or increased high-intensity exercise. Only 13 (34%) of these women reported changes in time bleeding compared to 36 of the 53 women (68%) who either decreased or did not do high-intensity exercise (p=0.003).

Most participants (75.0%) were not using any form of hormonal contraception. Over half of the total sample (52.6%) indicated they experienced a change in their cycle length during lock down. A quarter of the sample (25.3%) experienced increases in cycle length, and∼20% experienced a decrease. A change in bleeding time was reported by 267 (36%) participants. Of the symptoms listed in figure 2, decision tree analysis (Supplementary file, Figure 4) suggested that use of hormonal contraception was the biggest contributing factor in whether cycle length had changed. Amongst the 558 of women that were not using hormonal contraception, 58% noticed a change in cycle length, compared to 41% of the 187 who did use hormonal contraception (Table 3), which was a significant difference (p<.001).

## 4. Discussion

The purpose of this research was to identify if exercising females had experienced a change in their menstrual cycle throughout the COVID-19 pandemic lockdown period. Seven-hundred and forty-nine participant responses were included for final analysis, over half of which cited a change in their menstrual cycle. Over one third of participants experienced a change in bleeding patterns (35.6%). Secondly, the study aimed to identify the main contributing factors to such changes in the menstrual cycle. The greatest contributing factor to reported changes was stress. Some of the symptoms exacerbated by stress through the COVID-19 pandemic can be off-set by increases in fruit and vegetable intake and performing high intensity exercise. This study is the first to detail the implications of the pandemic on the menstrual cycle of females. The data provided herein not only highlights the importance of stress in relation to maintaining a consistent menstrual cycle but also provides important detail for practitioners/clinicians when managing patients with irregularities of the menstrual cycle.

Most menstrual cycle-associated symptoms that showed evidence of change during the COVID-19 pandemic were psychosocial. Over half of all participants reported a change in mood (54%), irritability (59.9%), emotional feeling (66.5%), worry (61.0%), feeling distracted (56.6%), lack of concentration (56.9%), motivation (62.3%) and focus (55.7%). These reported symptoms were consistent irrespective of whether the participant used a hormonal contraceptive or whether cycle length had changed. Most Hormonal contraceptives provide synthetic hormones to the body and regulate the menstrual cycle through synthetic pathways (Elliott-Sale and Hicks, 2018), and are also associated with various symptoms and side effects (Martin *et al*., 2017; Vitzthum and Ringheim, 2005; Nault *et al*., 2013). The data here has shown that whilst the cycle length of a hormonal contraceptive user may not change as readily in relation to the external environment, symptoms and side effects may still be present and can be affected by environmental changes.

There were some differences in the symptoms reported between elite and sub-elite athletes, compared to non-elite respondents. Fifty-four percent of elite female athletes indicated that bloating was a symptom they had increasingly experienced during COVID-19 lockdown, which was significantly higher than the reported prevalence of bloating amongst the other participants. This proportion is higher than previously reported amongst elite athletes populations (Martin *et al*., 2017), suggesting the COVID-19 lockdown period is likely to have contributed to this higher prevalence. Shunting of blood during exercise, away from the gastrointestinal (GI) tract towards the muscles, may contribute to increased GI stress in the form of bloating (ter Steege and Kolkman, 2012; Rehrer and Meijer, 1991) and could provide explanation toward the increases in bloating experienced by elite athletes, considering the higher levels of exercise they perform, and the increases in aerobic activity reported by 40% of participants. Despite the differences between elite and non-elite athletes, there was a clear overall change in bloating for all participants, which aligns with increasing reports of the effects of stress and anxiety on the gut (Wilson, 2020; Whitehead *et al*., 1992; Simpson and Stakes, 1987). It was also reported that higher level competitors may have been less likely than other women to experience reductions in; sex drive, focus and motivation and increases in; constipation, feeling emotional and lower back pain, compared to non-competitive athletes, however this was not significant. Whilst elite sport competition has been cancelled or postponed during the COVID-19 pandemic lockdown, it is likely that these athletes will have access to various additional support networks (Bowes, Lomax and Piasecki, 2020) and thus are able to manage rapidly changing environments more so than their non-elite counterparts.

Influences of nutritional intake and exercise on the menstrual cycle have been well documented (Torstveit and Sundgot-Borgen, 2005; Hashim *et al*., 2019; De Souza *et al*., 2017; Brown and Brown, 2010). During the past six months, across the globe, varying degrees of lockdown have arisen, inevitably affecting the ability of many females to carry out exercise. All sporting facilities were closed, and in some countries, restrictions have been limited people to within a 1 km radius from their homes. During such a climate, nutritional habits have inevitably changed (Di Renzo *et al*., 2020; Rodríguez-Pérez *et al*., 2020; Pellegrini *et al*., 2020). Our data report similar dietary changes. Not unexpectedly participants reported to have increased alcohol intake, consumption of cooked and baked goods and to have generally increased consumption of unhealthy foods, which is not unsurprising given the evidence of comfort eating in heighted stress (Roberts, Campbell and Troop, 2014; Klatzkin *et al*., 2019). Interestingly, an increase in dairy assumption was associated with cycle length changes seen in hormonal contraceptive users only. This is somewhat of an unexpected finding given the regularity of menstrual cycles that oral contraceptives provide (Elliott-Sale and Hicks, 2018), although there is some possible differences in efficacy and absorption between individuals taking oral contraceptives (Jung-Hoffman and Kuhl, 1990), which could possibly explain why some oral contraceptive pill users here still experienced cycle length changes. However, as we have no detailed pharmacodynamics data associated with the questionnaire this cannot be validated within this study.

The greatest contributor towards changes in symptoms was stress. There was a significantly greater increase in symptoms when participants reported high levels of worry about personal and family health. Similarly, bleeding time was more likely to have changed if high level of stress regarding job security was reported. Interestingly, fruit and vegetable intake were able to off-set some changes in cycle length associated with stress, and high intensity training was able to regulate stress regarding job security. Clearly, the current climate surrounding COVID-19 has been a stressful event for many (López *et al*., 2020; Rehman *et al*., 2020; Odriozola-González *et al*., 2020). This data highlights the great impact that such stress can have on female physiology, not solely on mental health and well-being. Current education around such possible influencers on the menstrual cycle is quite limited amongst the female population (Ayoola, Zandee and Adams, 2016; Larsen *et al*., 2020).

Given that the current climate is likely to continue for the foreseeable future, it is important that females become aware of the influence that stress will have on the menstrual cycle and are able to manage/control these changes to the best of their ability. It is deemed that long-term stress could have a significant negative impact on fertility and female reproductive health, (Palomba *et al*., 2018; Negro-Vilar, 1993; Sominsky *et al*., 2017), as well as the suggestion that exacerbated stress can induce premature brain ageing (Prokai and Berga, 2016). Therefore, it is especially important for females of childbearing age, and clinicians alike, to consider a number of possible factors, including stress, that could be contributing to any presenting changes in the menstrual cycle. Interestingly, exercise and a balanced sound nutritional intake were able to off-set some of the heightened stress, which aligns with what is already recommended for dealing with extreme stressors (Chan *et al*., 2019; Paolucci *et al*., 2018; Costigan *et al*., 2016; Ramón-Arbués *et al*., 2019; Owen and Corfe, 2017).

As with all research, there are limitations with this study that should be acknowledged. The data herein comes from a range of countries in which there were a range of rules and regulations, and therefore experiences, with regards to lockdown, therefore possibly influencing the participants differently. However, the countries assessed in the study all documented some degree of change in lifestyles and working environments. Our sample also included hormonal contraceptive users as well as non-users. The regularity of cycles when utilising combined oral contraceptives, due to exogenous hormones that they contain, is well documented, thus including such participants may limit the numbers of females that have seen cycle length changes. However, it was important not to exclude such females as it is known 34 % of the general population (Cea-Soriano *et al*., 2014) and around 50% of elite athletes (Martin *et al*., 2017) use such methods. Stress may not just change cycle length, it can also contribute to symptoms experienced which we were able to document in both hormonal contraceptive users and non-users. Finally, our data only provides a small insight into the changes experienced by females during the COVID-19 pandemic and does not provide any longitudinal data that could document the rise and fall of symptom exacerbation or the degree of severity that the females may have been affected by such environmental changes.

## 5. Conclusions

In conclusion, lifestyle changes experienced by females throughout the COVID-19 pandemic lockdown period can impact menstrual cycle length and symptoms. Over half of all females reported a change in their cycle length and over a third experienced changes in bleeding patterns. The most substantial changes arise in psychosocial symptoms, which is somewhat expected given previously reported links between the pandemic lockdown period and mental health. The stress caused by the pandemic is the most significant contributor to menstrual cycle changes. This is important and highlights the potential for long-term stress to influence female fertility and other health consequences. With the current uncertainty of the COVID-19 pandemic, and a possibility of a second wave during the northern hemisphere winter season, menstrual cycle changes could become a serious consequence. It is therefore important to educate females about the influence of stress as well as to ensure clinicians are fully aware of the reasons why menstrual cycle changes may arise.

## Supporting information

Supplementary file 1. Decision tree analysis

Supplemental file 2. Questionnaire

## Data Availability

The data that support the findings of this study are available from the corresponding author, upon reasonable request

## Supplementary Materials

The following are available online. Supplementary File 1: Figure 4; Decision Tree Analysis. Supplementary File 2. Questionnaire.

## Acknowledgments

The authors would like to that all participants for taking the time to participate in this study, without which such research would not be possible.

## Data availability statement

The data that support the findings of this study are available from the corresponding author (JP), upon reasonable request.

