## Supplementary figures and images for "How lifestyle changes within the COVID-19 global pandemic have affected the pattern and symptoms of the menstrual cycle"

### Supplementary file 1. Decision tree analysis

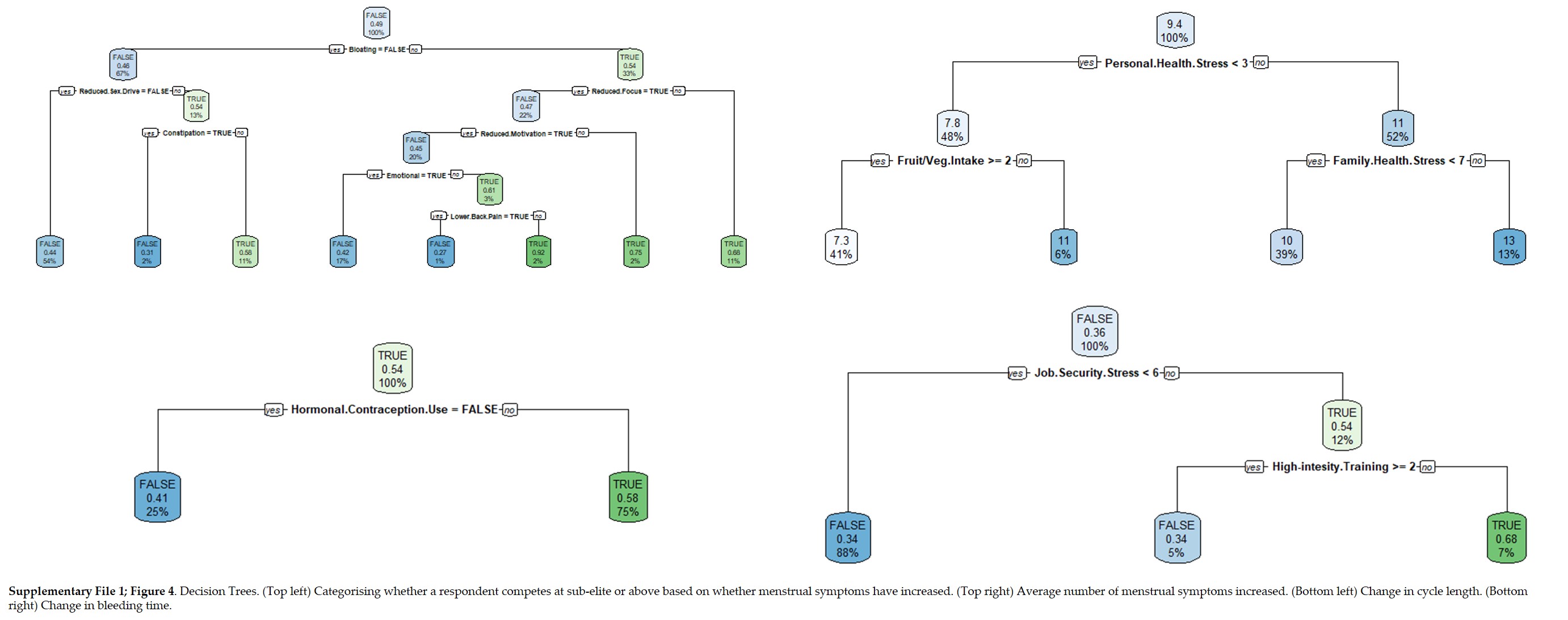
