## Supplemental file 2. Questionnaire for "How lifestyle changes within the COVID-19 global pandemic have affected the pattern and symptoms of the menstrual cycle"

*Investigating the impact of COVID 19 on menstrual cycle symptoms and side effects*

*Participant ID Number:*

Please select a participant ID number using 2 letters and 3 numbers. This will be used to store your data anonymously. Please make a note of this if you later wish to withdraw your answers.

*Introduction*

*Each question in this section will have an open box for the participant to fill in their answers. For question 4 and 5 there will be a number of possible selections for the answer, as listed below.*

1. Age; 18-24; 25-29, 30-35, 36-40, 41-45, 45+
2. Occupation
3. Primary Sport
4. Level of competition: Active/fitness training (less than 5 hours/week), sub elite (training towards a specific competition goal, 5-8 hours/week), elite training (national and international competition, 8+ hours/week, professional athlete)
5. Menstrual status; Are you using any form of contraception Y/N. If yes please state what type.
6. If yes to Q5 please state what type of contraception used. If N to Q5 would you describe your cycles as regular, irregular or totally absent?
7. Prior to COVID what is, roughly, the number of days between day 1 of bleeding and the onset of your next cycle?
8. Martial status; single, married, divorced, widowed, living with partner, other (please state)
9. Ethnicity; White, mixed/multiple ethnic groups, Asian/Asian British, Black/African/ Caribbean/Black British, Other ethnic groups (please describe), prefer not to say

*Working status*

10. Prior to COVID19 were you working/studying; full time, part time, job share, training as an athlete full time, unemployed, carer, other (please specify).
11. During the COVID19 pandemic how are you now working/studying; nothing has changed, full time at home, part time at home, furloughed 100% pay, furlough at less than 100% pay, other (please state)
12. Do you have children? Y/N If yes proceed to Q11 if no move to Q15
13. Please tick the appropriate box(es) for the age of your children: <3, 3-6, 6-10, 10-14, 14-18, 18+
14. Do one or more of your children require home-schooling and/or other support during the COVID19? Y/N

Please elaborate if necessary

15. Do you children affect your normal working routine Y/N
16. On a scale of 1-7 please highlight how this affects your daily stress levels (1 being not at all, 7 being extremely stressful)
17. On a scale of 1-7 please highlight how worried you are about your job security during this COVID pandemic and going forward from here (1 being not at all, 7 being extremely worried)
18. On a scale of 1-7 please highlight how worried you are about personal finances during this COVID pandemic (1 being not at all, 7 being extremely worried)

*Training*

19. How has the amount of exercise you perform changed during the lockdown period compared to before the lockdown? (*tick one box only for each activity*)

| Exercise activity | 1  About the same | 2  Increased | 3  Decreased | 4  I never do this |
| --- | --- | --- | --- | --- |
| Sports-specific skill training (ie practice of skills associated with your sport) |  |  |  |  |
| Aerobic endurance training (i.e. continuous running/cycling/rowing) |  |  |  |  |
| Aerobic-based interval training (i.e. repetitions >2 min duration, short recoveries) |  |  |  |  |
| High-intensity interval and sprint training (i.e. repetitions <2 min duration, long recoveries) |  |  |  |  |
| Multi-joint resistance training (i.e. barbell, dumbell, kettlebell, machine exercises) |  |  |  |  |
| Single-joint resistance training (e.g. calf raises, theraband exercises, lateral raise) |  |  |  |  |
| Plyometrics (i.e. jumping, hopping, skipping, bounding) |  |  |  |  |
| Body weight conditioning (e.g. press-ups, burpees, high knees, star jumps) |  |  |  |  |
| Core stability/pilates (exercises specifically for the trunk/abdominal region) |  |  |  |  |
| Static stretching |  |  |  |  |

*Menstrual experiences*

20. How have the symptoms you experience during your menstrual cycle changed, if at all, during the lock down.

| Menstrual cycle symptom | 1  About the same | 2  Increased | 3  Decreased | 4  I do not experience this symptom |
| --- | --- | --- | --- | --- |
| Lower back pain |  |  |  |  |
| Headache/migraines |  |  |  |  |
| Nausea/vomiting |  |  |  |  |
| Diarrhoea  Constipation  Dizziness/light headed. |  |  |  |  |
| Bloating |  |  |  |  |
| Weight gain |  |  |  |  |
| Water retention |  |  |  |  |
| Tiredness/Fatigue |  |  |  |  |
| Lack of coordination |  |  |  |  |
| Sleep disturbances |  |  |  |  |
| Hot flushes/ Sweating |  |  |  |  |
| Breast pain/tenderness |  |  |  |  |
| Cravings |  |  |  |  |
| Reduced appetite |  |  |  |  |
| Flooding/Leaking |  |  |  |  |
| Blood clots |  |  |  |  |
| Mood changes/Mood swings |  |  |  |  |
| Feeling irritable/angry |  |  |  |  |
| Feeling emotional |  |  |  |  |
| Feeling worried/ anxious |  |  |  |  |
| Feeling distracted |  |  |  |  |
| Reduced ability to concentrate |  |  |  |  |
| Lack of motivation |  |  |  |  |
| Lack of focus |  |  |  |  |
| Reduced sex drive |  |  |  |  |
| Increased sex drive |  |  |  |  |

21. During the lockdown has the length of your cycle (number of days between the first day of your period (day one of bleeding) and the first day of your next period (day one of bleeding)); increased, decreased or stayed the same
22. If you stated your cycle has changed in Q21 please state how this has changed. For example My cycles have got longer/shorter or they have become absent during lockdown.
23. Has the length of time of bleeding during your period; increased, decreased, stayed the same?
24. During the lockdown, how would you describe your menstrual blood loss? (same, increased, decreased)
25. Only answer this question if you stated an increase or decrease to Q21, 23 or 24.

If your cycle symptoms and/or length has changed please elaborate on how you have managed these changes for example, I exercised more to deal with my symptoms, or I have tried to eat different foods to help my symptoms or I have just dealt with them and not changed my routine. Please describe.

26. If your cycle symptoms and/or cycle length has changed, please rate on a scale of 1 – 7 how stressed you feel about these changes (1 being not at all, 7 being extremely stressed)

*General Health*

27. On a scale of 1-7 (1 being not worried, 7 being extremely worried) please rate how worried you are about your own health during this pandemic.
28. On a scale of 1-7 (1 being not worried, 7 being extremely worried) please rate how worried you are about your families health during this pandemic.
29. Have you been diagnosed with COVID through diagnostic tests during the lock down period? Y/N
30. Have you had symptoms and suspected you have had COVID therefore had to self isolate during this lock down period? Y/N
31. Has anyone in your household had symptoms of the virus and therefore you have had to self-isolate? Y/N
32. In terms of dietary patterns please select the most appropriate selections for your nutritional intake during this lockdown/pandemic period.

| Nutritional food group | 1  About the same | 2  Increased | 3  Decreased | 4  I never do this |
| --- | --- | --- | --- | --- |
| Eating fruits and vegetables |  |  |  |  |
| Eating dairy products |  |  |  |  |
| Eating red meat e.g. beef, lamb, pork (protein) |  |  |  |  |
| Eating white meat e.g. chicken, turkey |  |  |  |  |
| Eating processed meat e.g. bacon, sausages, ham |  |  |  |  |
| Eating fish |  |  |  |  |
| Eating grains/carbohydrates (for example rice, pasta, potatoes) |  |  |  |  |
| Eating meat alternatives (quorn, tofu) |  |  |  |  |
| Eating healthy fats (nuts, seeds, avocados, nut butters, oils) |  |  |  |  |
| Eating less healthy/processed foods (cakes, sweets, biscuits, chocolates) |  |  |  |  |
| Eating more home baked/cooked foods |  |  |  |  |
| Eating more take outs |  |  |  |  |
| Drinking alcohol |  |  |  |  |

33. Please feel free to add anything else you feel is relevant to our questionnaire
